# Beyond ‘Expressed Emotion’: Development and validation of a contextual measure of family communication and involvement in the context of psychosis in Ethiopia

**DOI:** 10.1101/2025.11.05.25339558

**Authors:** Dessalegn Kiross, Denise Taylor, Kassahun Habtamu, Wubalem Fekadu, Atalay Alem, Mekonnen Tsehay, Eleni Misganaw, Crick Lund, Tessa Roberts, Ashok Malla, Alex Cohen, Kia-Chong Chua, Tosin Popoola, Charlotte Hanlon

## Abstract

**Background and Aim:** Family caregiving environments significantly influence psychosis outcomes, yet existing measures like Expressed Emotion lack cultural relevance in non-Western settings. This study aimed to develop and validate a socio-culturally appropriate Family Communication and Involvement (FCI) scale in Ethiopia.

**Methods:** An initial FCI scale (version 0) was generated by integrating items from existing measures of aspects of family communication with findings from an ethnographic study with Ethiopian families caring for a relative with psychosis. To ensure content validity and clarity, items underwent translation, back-translation, expert consensus review, and cognitive interviews (n=20). The resulting FCI v1.0 (37 items) was piloted with 201 people with psychosis in rural Ethiopia. Exploratory factor analysis, item-item/item-total correlations, and test-retest reliability (n=50) informed stakeholder discussions to reduce items, leading to a 24-item, two-factor structure. The final 24-item scale (FCI v2) was validated with a separate sample of 401 people with psychosis in Addis Ababa. Confirmatory factor analysis (CFA) assessed structural validity. Convergent validity was evaluated against the WHO Disability Assessment Schedule and the Brief Psychiatric Rating Scale–Expanded.

**Results:** The scale demonstrated robust psychometric performance: excellent internal consistency (Cronbach’s α = 0.92) and acceptable convergent validity, correlating with disability (WHODAS, r = - 0.45) and symptom severity (BPRS-E, r = -0.48). Confirmatory factor analysis supported the 24-item bi-factor with acceptable model fit.

**Conclusion:** The FCI scale is a culturally grounded, psychometrically sound tool for assessing family communication and involvement in Ethiopia. It holds promise for use in similar non-Western settings where family support and sociocultural influences shape mental health care.

## Background

Psychotic disorders, such as schizophrenia or affective psychoses, demand comprehensive treatment strategies that extend beyond symptom management to address the broader psychosocial environment^1^. Family relationships and caregiver behaviours are pivotal factors influencing recovery trajectories, symptom recurrence, and quality of life for people with psychosis ^2^. High-quality and highly replicated evidence supports the effectiveness of family interventions in managing psychosis ^3 4^. Recognizing this, guidelines from high-income countries, as well as recent recommendations from the World Health Organization (WHO), emphasize the inclusion of family interventions as an essential component of psychosis care^1 5^.

A substantial body of research highlights the profound impact of familial attitudes and behaviours on the clinical outcomes of people with psychosis^6 7^. The construct of Expressed Emotion (EE), originating in Western studies, has been instrumental in identifying patterns of negative family interactions, such as criticism, hostility, and emotional over-involvement, that are strongly associated with relapse rates^8^. The Camberwell Family Interview (CFI), regarded as the gold-standard tool for assessing EE, categorizes family environments as high or low EE, providing insight into the influence of family interactions on illness progression. The EE-relapse link has been replicated across numerous Western and non-Western settings^6 7 9 10^ and led to the development of several effective family interventions that target reducing EE^4 11^.

Despite its widespread use in the measurement of EE, the CFI presents several challenges as a tool ^12–14^. First, CFI is resource-intensive, requiring extensive training and several hours for administration, which limits its feasibility in routine clinical practice, particularly in low-resource settings. Second, it predominantly focuses on negative family interactions, overlooking supportive or protective aspects of family dynamics that may play a crucial role in an individual’s recovery journey ^15 16^. Third, it adopts a unidirectional perspective, assessing family interactions based on observation of family behaviour, and failing to capture the perspectives of people with psychosis, despite the inherently reciprocal nature of family relationships^17^. Fourth, cross-cultural research has questioned the validity of the EE construct itself, particularly the emotional overinvolvement index. Behaviours considered as high EE, such as intense worry and close monitoring, were reported as signs of compassion and support in some non-Western settings^12 18^

Recognizing these limitations, efforts have been made to develop alternative, brief measures of family interaction in the context of psychosis^19–22^. However, many of these tools remain anchored in Western frameworks and may not adequately capture the cultural specificities of family life in diverse settings^16 17^. Notably, no validated measures have been developed in non-Western LMICs that reflect lived experiences in these contexts. This creates a critical gap in both research and clinical practice, limiting the evaluation of effective family-based interventions for psychosis^13 23^.

To address this gap, this study aimed to develop and validate the Family Communication and Involvement (FCI) scale, a contextualized measure grounded in ethnographic research.

## Methods

This study followed the standard guidelines for the development and validation of patient-reported outcome measures (PROMs)^24^, encompassing item development, scale development, and psychometric evaluation. To achieve a culturally sensitive and valid measure of family communication, we conducted an exploratory sequential mixed-methods study (Figure 1).

### Phase 1: Item development

#### 1.1. Ethnography

A focused ethnography was conducted to explore family communication and involvement in Ethiopian households with a person with psychosis, forming the basis for item development. The study included 14 families, five urban from Addis Ababa, and nine rural from the Sodo district, south-central Ethiopia. Rural participants were recruited from the PRIME^25^ Study database through a local gatekeeper, while urban participants were identified via the family health team of the health centre serving a district. Households were purposively selected to ensure diversity in illness severity, gender, socioeconomic status, and treatment status.

Data collection involved a minimum of 30 hours of observation per family and semi-structured interviews with stakeholders, including community leaders, mental health professionals, people with psychosis, and family caregivers. The integration of observations and interviews aimed to provide a socially embedded understanding of family communication, caregiving practices, and the broader context of mental health care in Ethiopia. Findings from the ethnography were used to develop an initial pool of items reflecting contextually relevant aspects of family communication and involvement.

##### 1.2. Review of Existing Scales

A focused literature search was conducted to identify potentially relevant scales used to measure aspects of family interaction in the context of psychosis globally. These scales were evaluated based on their item development process, theoretical linkage to the EE construct and other aspects of family interaction, and their reported psychometric properties.

Concepts derived from the ethnography were systematically mapped onto items from the identified existing scales to assess conceptual overlap and identify aspects of communication or involvement that were not captured by existing scales.

##### 1.3. Expert consensus meetings

A panel comprising nine international and 13 local experts (psychometricians, psychiatrists, clinical psychologists, family caregivers, people with psychosis) was convened through online and in-person meetings.

The initial consensus meetings were used to: (1) review the ethnography findings to decide whether existing measures could be adapted or if a new scale was needed, and (2) evaluate an initial pool of potential items (derived from both existing scales and ethnographic insights) for comprehensiveness and content validity. The item generation and initial review process was conducted in English.

### Phase 2: Scale development and pilot testing

#### 2.1 Cognitive interviews

Cognitive interviewing^26^, a technique to evaluate participant understanding and response processes for survey items, was employed. The first author (DK) conducted the interviews using the initial draft scale (translated into Amharic) with 20 people with psychosis attending the outpatient mental health clinic at Butajira General Hospital. This hospital operates an outpatient mental health clinic and serves as a research site for large epidemiological studies of psychosis in Ethiopia^27 28^. People from across the district access mental health services at this clinic. Participants were purposively selected to represent diverse education levels and urban/rural residences. Eligibility criteria included a diagnosis of psychosis for at least six months, age 18 years or older, co-residence with a family caregiver, capacity to consent, and ability to converse in Amharic. Written informed consent was obtained from all participants before their involvement. The interview was conducted in Amharic (the local official language).

During interviews, participants were asked to explain their interpretation of each question, paraphrase questions in their own words, and provide their reasoning for their responses. This process was designed to identify issues related to comprehension, clarity, wording sensitivity, and the appropriateness of response options.

#### 2.2. Second Expert Group meeting

Following the cognitive interviews, the findings were presented to the same expert consensus group convened in Phase 1, leading to modifications to item wording, formatting, and the response categories. Consensus was obtained on a revised version of the scale (FCI-vl) suitable for pilot testing.

#### 2.3. Pilot testing

The pilot study aimed to investigate the factor structure, item-item and item-total correlations, and test-retest reliability of the draft FCI-v1 scale.

##### Sample size and recruitment procedure

The pilot study was conducted in the Gurage zone (Sodo and Butajira districts), south-central Ethiopia. The sample size for the FCI scale development was determined based on guidelines for factor analysis. While there is no definitive consensus, recommendations generally suggest a participant-to-variable ratio of 5:1 to 10:1, with a minimum total sample size of 200^29–31^. To align with these recommendations, a total of 201 participants were recruited. A randomly selected subsample (n = 50) underwent retesting one week later to assess test-retest reliability.

Participants were recruited from both community and hospital settings. The inclusion criteria were the same as the criteria for the cognitive interviews described above. Data collectors used the PRIME^25^ and the Butajira cohort databases^28^ to identify participants from the community. Hospital participants were recruited from the outpatient mental health clinics of Butajira and Sodo primary hospitals.

For hospital-based recruitment, clinicians assessed the capacity to consent. A clinic manager then linked interested participants to the data collectors. For community -based recruitment, research assistants familiar with the area helped identify and locate potential participants. Mental capacity was not assessed clinically for community participants, as data collectors were not mental health professionals. However, only those who were not acutely ill, could converse in Amharic, and were able to provide written informed consent, were included. Written informed consent was obtained from all participants before their involvement. Data was collected via interviewer -administered questionnaires in Amharic (the local official language).

##### Data collection procedures

The pilot study was conducted from March 1 to April 20, 2023. Ten experienced data collectors from the district were trained for three days by the first author (DK). Data were collected using the Open Data Kit (ODK) software^32^.

The FCI-vl was administered to participants alongside standardized sociodemographic and clinical assessment interviews. Respondents used a 4-point Likert scale (0-3) to indicate the frequency of each item: “all the time” (3), “most of the time” (2), “rarely” (1), and “none” (0). To simplify interpretation, items 22-37 were reverse-scored. Total scores ranged from 0 to 111, with higher scores indicating helpful family communication.

##### Data analysis

Descriptive statistics (frequency, percentage, mean, and standard deviation) were used to examine the distribution of responses to each item. Items with very low mean values compared to other items were considered for revision or deletion based on expert consensus.

We used polychoric correlation(r) to examine the correlation of items within the scale ^33^. Item-scale and item-item correlations were calculated to assess the correlations between items and between an item and the total scale score^34^. We considered significant floor or ceiling effects when over 15% of the participants achieved the lower or maximum score for the scale. We used Cronbach’s alpha (α) to determine the internal consistency of the scale^35^.

Exploratory factor analysis (EFA) was conducted using the Diagonally Weighted Least Squares (DWLS) estimator and oblimin rotation method, to investigate the factor structure of the scale ^36^. We evaluated the multivariate normality of the data through visual examination of a Q-Q plot and analyzing kurtosis and skewness. Then, we confirmed that the correlation matrix was factorable with Bartlett’s test of sphericity and the Kaiser-Meyer-Olkin (KMO) Measure of Sampling Adequacy (MSA)^37^. The number of factors to be extracted was decided using a combination of parallel analysis (Horn’s), scree plot, percentage of variance explained by each factor, and expert consensus.

The significance of factor loadings was determined using a power analysis approach (80% power, α = 0.05) to account for the inflated standard errors inherent to factor loadings compared to traditional correlation coefficients. To mitigate against Type I errors, stricter loading thresholds were applied based on sample size^37^. With our sample size of 201, a factor loading of 0.4 was considered significant. If an item loaded on more than one factor with >0.4, it was considered problematic. For factors to be considered independent, they had to have at least three items with significant loadings, internal consistency reliability of ≥0.70, and theoretical meaningfulness^37^. We used a two-way mixed effect Intra-Class Correlation Coefficient (ICC 3, k), absolute agreement to examine test-retest reliability^38^.

All analyses were conducted using R packages lavaan, psych, dynamic, and dplyr ^39 40^.

##### Expert consensus meeting

The initial EFA of the 37-item FCI-v1 identified several items that did not meet established criteria (items with factor loadings < 0.40 or cross-loadings > 0.40)^37^. An expert consensus meeting reviewed these findings, leading to the removal and revision of items, resulting in a 25-item FCI-v2. A subsequent EFA on FCI-v2 further refined the scale to 24 items, which was then administered in the validation study.

### Phase 3. Validation study

The validation study aimed to test the construct validity of the scale using confirmatory factor analysis and to evaluate convergent validity. Since there is no gold standard measure of family communication for the study population and setting, it was not possible to examine criterion validity.

We investigated convergent validity with symptom severity (measured using the BPRS-E^41 42^) and functional impairment measured using the WHODAS-V2^43^). The rationale was that the family caregiving environment plays a crucial role in shaping mental health outcomes, particularly in collectivist cultures where family support and conflict can significantly impact illness trajectories. Research has shown that dysfunctional family interactions exhibiting high levels of hostility, criticism, and over-involvement are associated with increased symptom severity and poorer functional outcomes in people with psychosis ^67^.

#### Study area, population, and data collection procedure

The validation study was conducted at Amanuel Mental Specialized Hospital (“Amanuel Hospital”) in Addis Ababa, Ethiopia. Amanuel Hospital is a tertiary referral hospital that serves the whole country. Data collection occurred between July and September 20, 2023. This study was set in a different location from the pilot study as the pilot study population was relatively small and stable, limiting the availability of people with psychosis for a larger validation sample.

Sample size determination and recruitment followed the same approach as the pilot study. A total of 401 individuals with psychosis were recruited, with 50 randomly selected for a test-retest reliability assessment after one week. Inclusion criteria remained consistent with those of the pilot study. Ten psychiatric nurses received a three-day training to conduct structured interviews.

The following instruments were administered, alongside a structured questionnaire used to collect data on sociodemographic and clinical information.

#### Family communication and involvement-version 2 (FCI-v2)

The FCI-v2 included 24 items; 13 (items 1-13) positively worded and 11 (items 14-24) negatively worded.

#### World Health Organization Disability Assessment Schedule (WHODAS-2.0)

The 12-item WHODAS-2.0 was used to measure the difficulty experienced by the person in performing daily activities and social participation in the past 30 days^44^. Each item is scored from 1 to 5. A higher score indicates severe difficulty. WHODAS has been validated in the Ethiopian context^45^.

#### Brief Psychiatric Rating Scale - Expanded version (BPRS-E)

The BPRS-E is a 24-item observer-rated symptom severity scale that indicates clinical symptom severity for psychosis, mood and somatic symptoms^41 42^. Each item is scored from 1 to 7. A higher score indicates severe symptoms. The BPRS-E has been used previously in the Ethiopian context .

#### Data analysis

Confirmatory factor analysis (CFA) was employed to test the two-factor structure of the FCI scale derived from the EFA findings from the pilot study. As is recommended for ordered categorical data, all latent variable models were estimated using the diagonally weighted least squares estimator with robust standard errors^36^. The goodness of fit was assessed with the following indices: χ^2^ test (p<0.05), Confirmatory Fit Index (CFI) >0.95, Tucker-Lewis Index (TLI)>0.95, and root mean square error of approximation (RMSEA)<O.Oδ^46^.

We used the Pearson’s correlation coefficient (r)^35^ to examine convergent validity with symptom severity (measured using the BPRS-E^41 42^) and functional impairment (measured using the WHODAS- V2^43^). Linear regression was used to determine the effect of the severity of symptoms and the level of disability of people with psychosis on family communication and involvement.

We used a two-way mixed effect Intra-Class Correlation Coefficient (ICC 3, *k*), absolute agreement to examine test-retest reliability^38^. All analysis were made in r using the r packages lavaan, psych, dynamic, and dplyr^39 40^.

### Ethical Considerations

This study received ethical approval from the Institutional Review Board of the College of Health Sciences at Addis Ababa University (Ref. 001/22/Psy). Written informed consent was obtained from all participants prior to their involvement in the study. The authors assert that all procedures contributing to this work comply with the ethical standards of the relevant national and institutional committees on human experimentation and with the Helsinki Declaration of 1975, as revised in 2013. All procedures involving human subjects/patients were approved by the Institutional Review Board of the College of Health Sciences at Addis Ababa University (Ref. 001/22/Psy).

## Results

### Phase 1 findings: item development

#### Ethnography findings

Culturally salient aspects of family interaction deemed essential for inclusion in a contextually relevant scale were identified (See Supplementary File 2, Table 1).

#### Item Mapping

The review identified 14 potentially relevant scales focused on family communication and involvement in the context of psychosis (8 caregiver-rated, 6 patient-rated) (see Supplementary File 1 for details).

#### Caregiver-rated scales

Camberwell Family Interview (CFI)^8^, Five-Minute Speech Sample (FMSS)^19^, Adjective Checklist (AC)^47^, Family Questionnaire (FQ)^22^, Family Attitude Scale (FAS)^48^, Family Emotional Involvement and Criticism Scale (FEICS)^49^, Patient Rejection Scale (PRS)^50^, Family Involvement Questionnaire (FIQ)- family version (Mohan and Thara, Personal communication).

#### Patient-rated scales

Assessment Of Expressed Emotion (AEE)^51^, Brief Dyadic Scale of Expressed Emotion (BDSEE) - Patient version^52^, Concise Chinese level of expressed emotion scale (CCLEE)^53^, Influential Relationships Questionnaire (IRQ)^54^, Level of expressed emotion scale (LEE)^20^, The Perceived Criticism (PC) Scale^55^.

Some concepts identified from the ethnography overlapped with existing items; however, several key aspects identified in the Ethiopian context, particularly practical aspects of interaction, were not adequately captured by existing scales (Supplementary file 2, Table 1).

#### Initial expert consensus outcomes and draft scale (FCI-v0)

The expert group meeting reached a consensus that it was necessary to develop a new scale that captures meaningful aspects of family interaction not represented in the existing scales, alongside relevant concepts from existing scales. This led to the creation of the initial draft scale, the Family Communication and Involvement Scale - Version 0 (FCI-v0). This version comprised 34 items (19 newly generated based on ethnography, 15 adapted from existing scales) with five -point Likert-type response options (‘all the time’, ‘most of the time’, ‘sometimes’, ‘very rarely’, ‘never’) (Supplementary file 2, Table 2).

### Phase 2 findings: scale development

#### Cognitive interview findings

Cognitive interviews (n=20) of the FCI-v0 (see supplementary file 2, Table 2) indicated that participants generally had no difficulty recalling events relevant to the items. However, several areas for improvement were identified:

- **Item clarity:** Restructuring Amharic items to place the frequency enquiry at the end of an item improved comprehension.
- **Response scale challenges:** The five-point Likert-type response categories proved problematic. Participants frequently selected the middle ‘sometimes’ option, potentially reflecting cultural norms discouraging direct negative evaluations of family. Reading the response options aloud with each item made it easier for participants to respond.
- **Wording:** Specific items were identified as difficult to understand, prompting the need for alternative wording to enhance clarity and comprehension (Supplementary File 2, Table 3).

#### Second expert group meeting outcomes and revised scale (FCI-v1)

The expert panel reviewed the cognitive interviewing findings and proposed several modifications:

- **Response scale revision:** A key recommendation was to remove the middle ‘sometimes’ response category, resulting in a four-point forced-choice format. This decision aimed to reduce potential neutral response bias.
- **Item refinement:** Items on medication administration against a person’s will were revised to specify different forms of coercion more clearly.
- **New item addition:** An item assessing the person’s perceived safety concern was added.
- **Wording and formatting:** Further iterative adjustments were made to item wording in both Amharic and English for clarity and cultural relevance (e.g., replacing less familiar concepts with local equivalents). Items were also reformatted from statements into questions to facilitate clearer and more intuitive responses. A detailed list of these modifications can be found in Supplementary File 2, Table 3.

The outcome of this process was the revised Family Communication and Involvement scale, Version 1 (FCI-v1), containing 37 items (21 positively worded, 16 negatively worded), ready for pilot testing (see Supplementary File 2, Table 4).

### Phase 3 findings - Pilot and Validation studies

#### 3.1.1. Characteristics of study participants

A total of 201 people with psychosis participated in the pilot study, and 401 participated in the validation study. The mean age of participants in the pilot study was 37.9 years (SD ± 13). Approximately one-third of these participants had never received formal education (Table 1). In the validation study, the mean age of participants was 37.6 years (SD ± 10.6). Just under a quarter (22.4%) of these participants had received higher education (Table 1). The participants in both studies had comparable durations of illness and time lived with a family caregiver.

**Table 1.** Socio-demographic characteristics of the pilot and validation study participants.

| Participant characteristics |  | Pilot study (n=201) |  | Validation study (n=401) |  |
| --- | --- | --- | --- | --- | --- |
|  |  | Frequency | % | Frequency | % |
| Sex | Female | 86 | 42.8 | 159 | 39.7 |
|  | Male | 115 | 57.2 | 242 | 60.3 |
| Residence | Urban | 91 | 45.3 | 340 | 84.8 |
|  | Rural | 110 | 54.7 | 61 | 15.2 |
| Education status | Unable to read/write | 46 | 22.9 | 11 | 2.7 |
|  | No formal education | 24 | 11.9 | 12 | 3.0 |
|  | Primary education | 86 | 42.8 | 151 | 37.7 |
|  | Secondary education | 36 | 17.9 | 137 | 34.2 |
|  | Higher education | 9 | 4.5 | 90 | 22.4 |
| Age category (years) | 18-25 | 44 | 22.0 | 46 | 11.5 |
|  | 26-35 | 60 | 29.8 | 138 | 34.4 |
|  | 36-45 | 44 | 22.0 | 130 | 32.4 |
|  | 46-55 | 31 | 15.4 | 60 | 15.0 |
|  | >56 | 22 | 10.8 | 27 | 6.7 |
|  |  | Mean | Standard deviation | Mean | Standard deviation |
| Age (years) |  | 37.9 | 13 | 37.6 | 10.6 |
| Duration of illness (years) |  | 11.0 | 8.3 | 11.4 | 8.6 |
| Duration of time lived with family (years) |  | 10.8 | 8.6 | 9.9 | 8.4 |

##### Item-level analysis of the pilot study data

No items were endorsed by everyone or not endorsed by anyone (Sup file 2, Table 5). Most of the items had item-total correlations that ranged from 0.60 to 0.78. Only item 24 (“In the past month, how much of the time was what you did supervised?”) and item 37 (“In the past month, how much of the time did your family push you to use alternative healing approaches to the medicine?”), both from the negatively worded items had an item-total correlation below 0.3 (Sup file 2, Table 5). The positively worded items all had an item-item correlation of >0.30, but not higher than 0.9. However, negatively worded items had a very low item-item correlation (<0.3) with the positively worded items and with each other. Four negatively worded items tapping into medication and help-seeking behaviour had a stronger correlation (items 30-33) with each other, ranging from 0.8 to 0.9 (supplementary file 2, Fig.1).

##### Exploratory factor analysis

An iterative EFA of the FCI-v1 scale was conducted on the pilot data to determine the underlying factor structure (see Supplementary File 2, Factor analysis- EFA). A stable three-factor structure emerged, explaining 61% of the total variance (Supplementary File 2, Table 7).

- Factor 1 consisted of 21 items, explaining 33% of the variance (Cronbach’s α = 0.95).
- Factor 2 consisted of 11 items, explaining 14% of the variance (Cronbach’s α = 0.82).
- Factor 3 consisted of 4 items, explaining 14% of the variance (Cronbach’s α = 0.92).

All item loadings exceeded 0.40, except for items 24 and 37. No items exhibited significant cross - loadings. Item-total correlation showed moderate to strong correlation for most items, except for item 37(Supplementary File 2, Table 8).

##### Expert group consensus for item reduction

Experts reviewed the pilot study findings and identified items targeting similar aspects of family dynamics within the draft scale. These items were merged. In addition, a few items were dropped from the scale due to their weak psychometric properties (Supplementary file 2, Table 9). This resulted in an intermediate 25-item scale (FCI-v2).

A subsequent EFA on this 25-item version indicated problems with the three-factor structure (Supplementary File 2, Fig. 4 and 5). Items 21 and 22, loading onto Factor 3, exhibited an excessively high correlation (r = 0.92). Item 22 also showed negative unique variance and multicollinearity, compromising factor stability (see Supplementary File 2, Factor analysis after item reduction for details). Both items originated from merged items related to treatment dynamics. Due to these psychometric issues, item 22 was removed.

The final Family Communication and Involvement Scale Version 2 (FCI-v2) comprised 24 items. EFA of the final 24-item FCI-v2 indicated a simplified two-factor structure explaining 55% of the total variance (Supplementary File 2, Tables 10-12). Correlations among the 13 positively worded items (Factor 1) ranged from 0.34 to 0.71. Correlations among the 11 negatively worded items (Factor 2) ranged from 0.17 to 0.71. Correlations between positively and negatively worded items ranged from 0.01 to 0.54 (Fig. 2).

#### Validation study

##### Descriptive Statistics

FCI-v2 was tested with 401 participants (see Table 1 for participant details). The average total score was 49.7 (±13.1), with scores ranging from 10 (lowest) to 72 (highest) across all participants (Supplementary file 2, Fig. 6).

Among the 24 questions:

- **Highest Score:** Item 21 (“Forced treatment”; reverse-scored) indicated low coercion (Table 2).
- **Lowest Score:** Item 2 (“Family discussions”) reflected infrequent communication (Table 2).

**Table 2.** Item descriptive statistics of the FCI-v3 scale in the validation sample (n=401)

|  |  | Item origin | Mean | Item-total correlation | Intra-class correlation (n=47) |
| --- | --- | --- | --- | --- | --- |
| In the past month, how much of the time does your family |  |  |  |  |  |
| 1 | Speak to you with a gentle voice? | IRQ | 2.11 | 0.58 | 0.74 |
| 2 | Talk things over with you? | IRQ | 1.48 | 0.48 | 0.81 |
| 3 | Show you a happy face? | IRQ | 2.15 | 0.68 | 0.66 |
| 4 | Sit with you? | BDSEE | 1.53 | 0.58 | 0.71 |
| 5 | Fulfil your needs within their capacity | ETHNO | 1.97 | 0.58 | 0.74 |
| 6 | Allow you to benefit from what the house can offer like other family members? | ETHNO | 2.14 | 0.56 | 0.69 |
| 7 | Make you feel safe living amongst them? | ETHNO | 2.19 | 0.65 | 0.79 |
| 8 | Genuinely listen to what you say? | ETHNO | 1.89 | 0.68 | 0.75 |
| 9 | Encourage you to do tasks without an aide? | ETHNO | 1.97 | 0.64 | 0.76 |
| 10 | Seem confident that you can accomplish a task without an aide? | ETHNO | 1.74 | 0.59 | 0.85 |
| 11 | Let you do what you want without any restriction | ETHNO | 1.74 | 0.65 | 0.71 |
| 12 | Involve you in beneficial tasks? | BDSEE/LEE/IRQ | 1.73 | 0.55 | 0.86 |
| 13 | Accept what you say as they do for other members of the family? | ETHNO | 1.75 | 0.72 | 0.67 |
| In the past month, how much of the time... |  |  |  |  |  |
| 14 | *Did you feel uncomfortable speaking openly about your feelings to family members? | ETHNO | 2.07 | 0.38 | 0.55 |
| 15 | *Did you feel excluded within the family? | ETHNO | 2.5 | 0.59 | 0.89 |
| 16 | *Did you yearn for someone in the family to talk to you about how you feel? | ETHNO | 2.19 | 0.57 | 0.56 |
| 17 | *Did your family interfere with what you did? | ETHNO | 1.96 | 0.50 | 0.69 |
| 18 | *Did you feel overcontrolled? | BDSEE | 1.97 | 0.46 | 0.83 |
| 19 | *Does your family consider you as a person who could not look after himself? | IRQ/LEE/BDSEE | 2.27 | 0.66 | 0.82 |
| 20 | *Do you feel used by your family because of your illness? | ETHNO | 2.65 | 0.58 | 0.77 |
| 21 | *Were you forced to use a treatment against your will? | ETHNO | 2.69 | 0.34 | 0.71 |
| 22 | *Does your family consider you as someone who does not have much to offer? | PRE | 2.31 | 0.63 | 0.88 |
| 23 | *Do disagreements occur because you aren't contributing to subsistence? | ETHNO | 2.43 | 0.62 | 0.62 |
| 24 | *Does your family get irritated over minor mistakes? | CCLEE | 2.18 | 0.68 | 0.73 |

ICC- intraclass correlation \*Reverse scored

The scale showed no floor effects (0% scored the minimum) and negligible ceiling effects (0.2% scored the maximum).

##### Confirmatory factor analysis

Bartlett’s test of sphericity, P < 0.001 and KMO (MSA = 0.92) confirmed the appropriateness of the correlation matrix for factor analysis. We tested the two-factor and bi-factor model of the FCI scale derived from the EFA (Table 3 and Figures 3 and 4).

**Table 3:** Factor loadings for the CFA models tested.

|  | 2-factor CFA | Bifactor CFA |
| --- | --- | --- |

| N = 401 |  |  | General Factor | Specific Factors |
| --- | --- | --- | --- | --- |
| Factor-1 | fci1 | 0.67 | 0.53 | 0.40 |
|  | fci2 | 0.59 | 0.33 | 0.61 |
|  | fci3 | 0.79 | 0.66 | 0.41 |
|  | fci4 | 0.69 | 0.45 | 0.59 |
|  | fci5 | 0.69 | 0.47 | 0.56 |
|  | fci6 | 0.67 | 0.50 | 0.45 |
|  | fci7 | 0.74 | 0.66 | 0.31 |
|  | fci8 | 0.79 | 0.69 | 0.36 |
|  | fci9 | 0.75 | 0.55 | 0.52 |
|  | fci10 | 0.69 | 0.49 | 0.51 |
|  | fci11 | 0.73 | 0.59 | 0.41 |
|  | fci12 | 0.63 | 0.44 | 0.49 |
|  | fci13 | 0.82 | 0.74 | 0.33 |
| Factor-2 | fci14 | 0.43 | 0.36 | 0.28 |
|  | fci15 | 0.78 | 0.68 | 0.39 |
|  | fci16 | 0.70 | 0.59 | 0.38 |
|  | fci17 | 0.61 | 0.41 | 0.67 |
|  | fci18 | 0.56 | 0.35 | 0.66 |
|  | fci19 | 0.79 | 0.69 | 0.38 |
|  | fci20 | 0.80 | 0.71 | 0.34 |
|  | fci22 | 0.45 | 0.41 | 0.16 |
|  | fci23 | 0.79 | 0.72 | 0.29 |
|  | fci24 | 0.78 | 0.72 | 0.24 |
|  | fci25 | 0.79 | 0.76 | 0.16 |
| Mean factor loadings | Factor 1 | 0.71 | - | 0.46 |
|  | Factor 2 | 0.68 | - | 0.36 |
|  | General factor | - | 0.56 | - |
| Factor correlation for 2-factor model | 0.67 |  | Na |  |

*\*All loadings are significant*

##### Model Fit indices

CFA demonstrated that both the two-factor and bifactor structures of the FCI scale showed acceptable fit, with the bifactor model providing superior fit indices (χ^2^: p < .001, SRMR 0.052, RMSEA 0.062, CFI 0.953) (supplementary file 2, Model fit). The bifactor model suggests that while two distinct dimensions of family communication and involvement exist, a broader underlying construct may also be present.

##### Convergent validity

Family communication and involvement scores were found to have a moderate correlation with symptom severity (r= -0.48, P< 0.001) and disability level (r= -0.45, P< 0.001) (supplementary file 2, Table 14).

Linear regression analysis indicated that both the severity of symptoms and the level of disability were associated with family communication. The BPRS coefficient indicated that for each one-unit increase in BPRS, the FCI score decreased by 0.28 units. The WHODAS coefficient indicated that for each one-unit increase in WHODAS, the FCI score decreased by 0.34 units (supplementary file 2, Table 15).

## Discussion

Research on the role of families in psychosis care in LMICs like Ethiopia is scarce due to the lack of culturally relevant measures of family communication. Existing instruments frequently fail to capture the complexity and nuances of family interactions in non-Western settings^12–14^. They are predominantly developed in Western contexts and often based on models like EE. The uncritical application of EE concepts and associated tools, such as the CFI, has questionable validity in contexts like Ethiopia ^12–14^.

Bhugra and McKenzie (2003) highlighted the challenges of applying EE universally, noting that cultural nuances significantly influence family interactions and emotional expression^13^. EE’s focus on overt criticism and emotional over-involvement can pathologize normative behaviours in collectivist societies. Furthermore, the CFI, often considered an EE gold standard, is limited by its primary focus on negative interactions and its reduction of complex family interactions into unidimensional “high” or “low” EE categories^12^.

To address this critical gap, we developed and validated the FCI scale that is suitable for the Ethiopian context. This tool was designed using a bottom-up approach to ensure it reflects the lived realities of Ethiopian families, filling a critical gap in the literature^13 23^. The scale’s development followed a rigorous, multi-stage process emphasizing cultural validity. An initial ethnography identified contextually relevant aspects of family communication, such as those not expressed verbally, and the significance of practical involvement in daily family routines. These aspects of interaction are not adequately represented in Western-centric models^8^. The insights from the ethnography informed the initial items of the scale. Subsequent expert consensus meetings refined these items to ensure they were clear, relevant, and contextually accurate.

A four-point forced-choice format was adopted to reduce neutral and acquiescent responses, based on evidence that such formats improve response quality^56 57^. Removing the middle ‘sometimes’ option helped minimize bias while preserving sensitivity to both positive and negative interactions. Psychometric testing confirmed the scale’s ability to measure both positive and negative interactions effectively, even in a cultural context where direct criticism is often avoided.

The FCI scale differs from traditional EE measures like the CFI by offering a more holistic view of family communication. It captures not only negative interactions but also positive ones, and also incorporates non-verbal dimensions identified as vital in the Ethiopian setting. This contrasts sharply with instruments that focus heavily on criticism^8^. Our findings also indicate an association between family communication measured by the FCI scale and the severity of psychosis symptoms and disability, aligning with broader literature on the role of family in psychosis care^6 58 59^. Future planned work aims to investigate the directionality of this association and determine whether specific aspects of family communication captured by the FCI predict patient outcomes^60^.

In terms of practical applications, the FCI scale can be useful for both clinical and research settings. Its brevity and clear structure facilitate ease of use, making it an efficient tool for assessing family communication and involvement in a way that informs potential interventions. While developed in Ethiopia, the scale can be useful in other LMIC contexts where families play a central role in caregiving and where challenges such as stigma, resource constraints, and reliance on traditional healers shape mental health care. By pinpointing specific communication areas that may benefit from intervention, the FCI scale can support clinicians in designing targeted strategies for improving family support. This aligns with existing literature that advocates for the inclusion of family-centered approaches in mental health care ^5 61^.

Beyond the FCI scale itself, the multi-stage, culturally-grounded methodological approach employed in this study offers a valuable template. The emphasis on initial ethnography to understand lived realities before item generation, followed by expert consensus and psychometric validation, represents a rigorous process for developing culturally-sensitive instruments. This bottom-up methodology could be effectively adapted for instrument development or adaptation in other settings where existing Western-centric measures may prove inadequate.

This study developed and validated a contextually relevant scale to assess family communication in psychosis care in Ethiopia. The FCI demonstrated strong reliability and validity, capturing important aspects of support, warmth, and practical involvement often overlooked by Western tools. By centering local caregiving practices, the scale supports Ethiopia’s shift toward community-based mental health care and underscores families’ therapeutic role in relapse prevention. Beyond its immediate context, the FCI demonstrates how culturally grounded methodologies can produce tools that respect local realities while offering insights for global mental health equity.

### Limitations

This study has few limitations. While the model generally fits the data reasonably well, the factor structure might benefit from further refinement. The Explained Common Variance in the EFA was 55%, which aligns with social science standards, where lower variance thresholds are common due to the complexity of human behavior^31^. While this is often considered acceptable in psychological research, future replications with larger, diverse samples could potentially improve this metric. Future research could also explore the FCI’s utility and validity in populations dealing with conditions such as severe depression, bipolar disorder, or even neurodevelopmental disorders, adapting items as necessary.

## Data Availability

All data produced in the present study are available upon reasonable request to the authors

## Acknowledgements

CH, AA, and EM receive support from the National Institute for Health and Care Research (NIHR) through the NIHR Global Health Research Group on Homelessness and Mental Health in Africa (NIHR134325), using UK aid from the UK Government. The views expressed in this publication are those of the authors and not necessarily those of the NIHR or the Department of Health and Social Care. CH also receives support from Wellcome Trust grant 223615/Z/21/Z.

For the purposes of open access, the author has applied a Creative Commons Attribution (CC BY) licence to any Accepted Author Manuscript version arising from this submission.

## Figure captions

*Figure 1. Phases of the scale development process (adapted from “Boateng, et.al., 2018, Best Practices for Developing and Validating Scales for Health, Social, and Behavioral Research: A Primer”)*

*Figure 2. Polychoric correlation matrix for FCI-v2, 24 items, n=201 *The size of the blue pie within the circle indicates the correlation strength*

*Figure 3.: Two-factor CFA model*

*Figure 4. A bifactor CFA model*

